# Prioritizing COVID-19 vaccination efforts and dose allocation within Madagascar

**DOI:** 10.1101/2021.08.23.21262463

**Authors:** Fidisoa Rasambainarivo, Tanjona Ramiadantsoa, Antso Raherinandrasana, Santatra Randrianarisoa, Benjamin L. Rice, Michelle V. Evans, Benjamin Roche, Fidiniaina Mamy Randriatsarafara, Amy Wesolowski, C. Jessica Metcalf

**Affiliations:** Department of Ecology and Evolutionary Biology, Princeton University, Princeton, NJ, USA; Mahaliana Labs SARL, Antananarivo, Madagascar; Department of Life Science, University of Fianarantsoa, Madagascar; Department of Mathematics, University of Fianarantsoa, Madagascar; MIVEGEC, Université de Montpellier, CNRS, IRD, Montpellier, France; Surveillance Unit, Ministry of Health of Madagascar; Faculty of Medicine, University of Antananarivo; Madagascar Health and Environmental Research (MAHERY), Maroantsetra, Madagascar; Direction of preventive Medicine, Ministry of Health of Madagascar; Department of Epidemiology, Johns Hopkins Bloomberg School of Public Health, Baltimore, MD, USA; Princeton School of Public and International Affairs, Princeton University, NJ, USA

## Abstract

**Background:** While mass COVID-19 vaccination programs are underway in high-income countries, limited availability of doses has resulted in few vaccines administered in low and middle income countries (LMICs). The COVID-19 Vaccines Global Access (COVAX) is a WHO-led initiative to promote vaccine access equity to LMICs and is providing many of the doses available in these settings. However, initial doses are limited and countries, such as Madagascar, need to develop prioritization schemes to maximize the benefits of vaccination with very limited supplies. There is some consensus that dose deployment should initially target health care workers, and those who are more vulnerable including older individuals. However, questions of geographic deployment remain, in particular associated with limits around vaccine access and delivery capacity in underserved communities, for example in rural areas that may also include substantial proportions of the population.

**Methods:** To address these questions, we developed a mathematical model of SARS-CoV-2 transmission dynamics and simulated various vaccination allocation strategies for Madagascar. Simulated strategies were based on a number of possible geographical prioritization schemes, testing sensitivity to initial susceptibility in the population, and evaluating the potential of tests for previous infection.

**Results:** Using cumulative deaths due to COVID-19 as the main outcome of interest, our results indicate that distributing the number of vaccine doses according to the number of elderly living in the region or according to the population size results in a greater reduction of mortality compared to distributing doses based on the reported number of cases and deaths. The benefits of vaccination strategies are diminished if the burden (and thus accumulated immunity) has been greatest in the most populous regions, but the overall strategy ranking remains comparable. If rapid tests for prior immunity may be swiftly and effectively delivered, there is potential for considerable gain in mortality averted, but considering delivery limitations modulates this.

**Conclusion:** At a subnational scale, our results support the strategy adopted by the COVAX initiative at a global scale.

## Introduction

The COVID-19 pandemic has resulted in a global health crisis resulting in an estimated 198 million cases and 4.2 million deaths (as of 31 July 2021) globally [1]. Until recently, non-pharmaceutical interventions, including social distancing, mask wearing, and travel restrictions were the primary mitigation measures. However, the development, approval, and distribution of several highly effective COVID-19 vaccines has resulted in a new era of public health response. The overall impact of mass vaccination on the global pandemic will depend on access to vaccines and ability to rapidly vaccinate populations [2]. There has been global competition to procure COVID-19 vaccines, and many low and middle income countries (LMICs) have been less successful than richer countries in securing vaccines [3] despite the COVAX initiative. COVAX is a World Health Organization (WHO) led initiative to promote vaccine access equity to LMICs and is set up to provide enough doses to immunize 20% of the population through distribution of multiple smaller batches [4]. Such efforts have provided essential baseline doses to LMICs, but even with vaccines in hand, countries face a number of logistical challenges. Some COVID-19 vaccines have extreme cold chain requirements and relatively short vaccine shelf-life that adds difficulties in avoiding vaccine wastage [4]. Likewise, achieving equity in vaccine coverage is always affected by heterogeneity in access to care [5] and may be further complicated by vaccine hesitancy [6]. However, many LMICs have more recent experience with mass vaccination campaigns (e.g., polio, measles) [7], which may provide an advantage in implementation relative to wealthier settings where mass vaccination campaigns have been less frequent in recent history.

Policy makers in LMICs face the central question of how COVID-19 vaccine doses should be allocated among populations in the face of these constraints and considerations of burden. So far, mathematical models developed to address the question of vaccine dose allocation have predominantly focused on the tradeoffs between prioritizing younger high contact individuals (which would reduce transmission) vs older high risk individuals (which would reduce mortality on infection). Previous work suggests that priority should be given to healthcare workers (HCW) and then to older adults, in line with rankings and guidelines provided by the WHO [8–10]. However, it should also be considered that focusing on distribution across age may neglect other drivers of inequity, such as geography and ethnicity [11]. For LMICs, a further important issue is local availability of personnel who can deliver doses, as numbers of HCWs may be limited.

Here, we explore the question of how regional vaccine dose distribution might be designed to minimize the burden of COVID-19 in Madagascar in the light of these features. We leverage data collected as part of a dashboard (www.covid19mg.org) that collates official reported cases of COVID-19 and census information. To date, Madagascar has officially reported a total of x cases and y deaths since March 20th 2020. Most cases (x) and deaths (y) are reported from the capital region of Analamanga, also the most populated region in the country with approximately 3 620 000 people representing 14% of the population. However, there is considerable uncertainty as to the burden of the disease on the Malagasy population to date, especially in rural areas where reporting rates are likely to be low and limited access to testing [12]. As prior infection by SARS-CoV-2 does generate immunity likely to be protective [13] against disease if not reinfection [14, 15], subnational variation in the trajectory of the pandemic to this point could influence the dose deployment strategy among regions that minimizes burden. However, inevitable uncertainties call for a focus on vaccine deployment efforts that are robust in the face of this underlying subnational variation rather than hinging on its characteristics.

On April 3rd 2021, Madagascar initiated the necessary steps to re-join COVAX and distributed the first doses of vaccines to healthcare workers and vulnerable populations on May 10th. As of July 20th 2021, 197 000 doses of an initial batch comprising 250 000 ChAdOx1-nCOV (Covishield™) vaccines were administered representing 0.73% of the national population (ourworldindata.org). These doses were distributed between the 22 different regions based on population size of each region. To inform the next steps in vaccine dose allocation strategies in Madagascar, we synthesize data on the regional distribution of elderly population and the number of reported cases and deaths in the country. Building from this background, we develop a mathematical model to investigate the optimal vaccine deployment strategy in the context of realistic constraints for mass vaccination campaigns based on health care worker availability, and known features of the burden of infection over age. We contrast four possible strategies for distribution based on a) population size, b) number of individuals over 60 years old, c) the number of reported cases, and d) the number of COVID-19 deaths by region. These strategies weigh different factors, i.e. those most at risk versus areas with the highest burden of the disease, and allow for a comparison of implementable strategies by weighing the overall number of deaths averted through each approach. Since deployment of vaccines to areas that had experienced large-scale outbreaks in the first waves of the infection might be less beneficial than deployment to less affected regions as a result of existing immunity in the population, we also evaluate sensitivity of our predictions to underlying susceptibility in the population, and explore the potential of rapid tests for seropositivity to guide vaccine distribution and avert mortality.

## Methods

### Data sources: demography, HCWs, and SARS-CoV-2 case distribution

Regional population size and age distribution were obtained from the 2018 census (Madagascar Institute of Statistics, INSTAT). The number of healthcare workers in each region was obtained from UNICEF database for Madagascar. SARS-CoV-2 cases and COVID-19 deaths were obtained from a dashboard (www.covid19mg.org) which compiles data communicated by the Ministry of Health on a daily basis. These data comprise PCR-confirmed cases, deaths per region as well as the number of tests performed nationally. Using these data, we ranked each region based on the size and the age distribution of the population and the number of healthcare workers in the region on the one hand and the situation with regards to the COVID-19 epidemic (officially reported cases and deaths) on the other hand.

### SARS-CoV-2 transmission model of Madagascar

We constructed an age-structured, stochastic SEAIR (susceptible, exposed, asymptomatic infection, symptomatic infection, and removed) transmission model by expanding previous work [16, 17] (see Supplementary figure S1). With this model, we simulated the trajectory of SARS-CoV-2 cases in each of the 22 regions of Madagascar under different assumptions about vaccination deployment among the regions (detailed below). For each region, the demography (age-structure and population size) was defined based on INSTAT statistics described above, while contact matrices were based on the social mixing patterns in the Mozambican population [18], since there is no contact matrix data available for Madagascar. We set R0 (the number of new infections per infectious individual in a completely susceptible population) to 2.5 as in [16] and simulated our model for a year. To quantify the burden of infection, we used the age-specific mortality risk (infection fatality rate by age) [19, 20], and each vaccination scenario was compared to a ‘no vaccination’ scenario. We explored a range of different starting proportions of the population susceptible, to reflect potentially varied histories of infection in each region. We initiated the outbreak by seeding 10 individuals in each region.

### Vaccination

To compare the impact of varying vaccine distribution among the 22 regions of Madagascar on total mortality, we assumed that the country received a single batch of COVID-19 vaccines that was sufficient to vaccinate 20% of the population. In our baseline scenario, we assume that 70% of those eligible for vaccination, regardless of age, will accept to be vaccinated with their full scheduled doses (Transparency International, unpublished). We initially do not assume that any information regarding previous infection status would be available, i.e. individuals who were previously infected may be vaccinated. The vaccine is assumed to work uniformly across age groups and be transmission and infection blocking with an efficacy of 76%, chosen to approximate the clinical vaccine efficacy against symptomatic infection seen for the ChAdOx1 nCoV-19 (AZD1222) [21, 22]. Finally, we assume that 50% of healthcare workers in each region would be mandated to vaccinate 20 people a day, based on experience of vaccination programs in Antananarivo. Vaccination follows an oldest-first strategy where vaccines are administered to individuals aged 60 years or older first. After all accepting individuals in the eligible group are vaccinated, individuals from the next (younger) age group are vaccinated and so on until all available doses are administered.

We then considered five allocation strategies of available doses:

1. Doses are distributed to regions uniformly (each region receives 4.5% of available vaccines)
2. Doses are distributed to regions based on population size (pro-rata),
3. Doses are allocated based on the distribution of people aged over 60 years between the regions (age),
4. Doses are distributed to regions based on the number of cases reported (cases),
5. Doses are distributed to regions based on the number of deaths reported (deaths)

The number of cases and deaths per region was obtained from the Madagascar COVID-19 dashboard (www.covid19mg.org) which collates the reported confirmed cases and deaths per region daily. For each allocation strategy, we then estimated the number of deaths and compared this value to a scenario without vaccination to calculate the number of averted deaths.

For each allocation strategy, we also varied the number of total doses available nationally (from 0 to 26 million covering 0-100% of the population), the vaccine acceptance rate (from 0-100%), the speed of rollout which is equivalent to the number of vaccinators per region (10-100% of healthcare workers in the region) and when the vaccination campaign began (0-200 days).

To investigate the effect of any potential existing immunity from prior infections, we considered two sets of initial conditions. In the first instance we assumed that 100% of the population was susceptible, all locations included at least 10 infected individuals, and the vaccination campaign would begin soon after the beginning of the simulation (within 10 days). In the second instance, we assumed that there is a baseline level of population-level immunity based on a uniform value for the entire country (0 - 40%) or proportional to reported cases (0-20%, see Supplementary Information).

To assess the benefit of targeting seronegative individuals through rapid testing, we included age-stratified seroprevalence and simulated different approaches to vaccine distribution.

## Results

Considering either population size, number of health-care workers (Figure 1A,B), number of reported cases, or numbers of confirmed deaths (Figure 1C,D) provides broadly similar overall priority rankings of regions (colours), with the region of Analamanga (AN, which contains the capital city, Antananarivo), consistently ranking highest, and the smaller, less densely populated regions (e.g., MK: Melaky) ranking lower.

**Figure 1:**
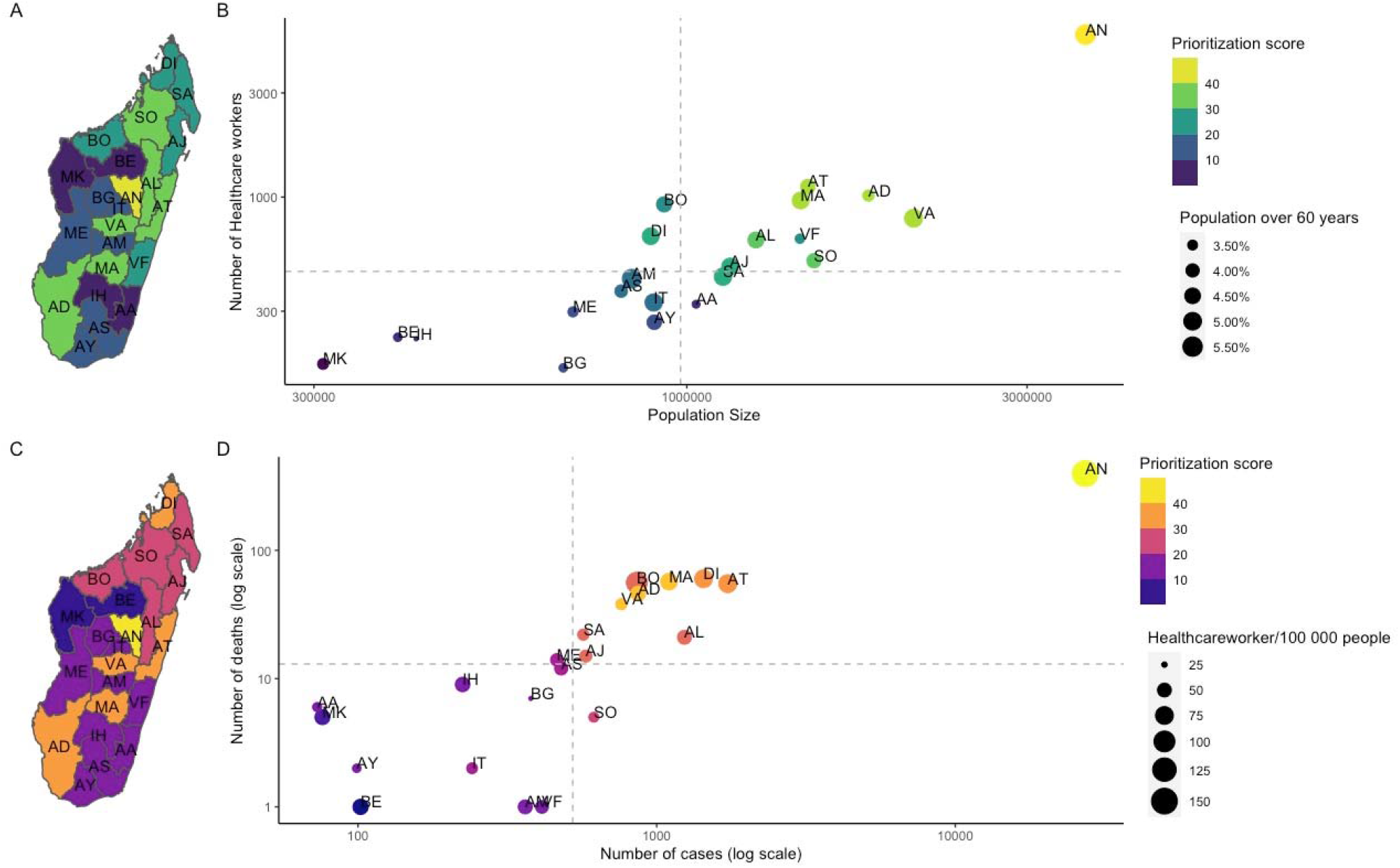
Demography, distribution of health care workers, SARS-CoV-2 cases, and deaths across Madagascar. A) The ranking for vaccine distribution based on the population size and number of healthcare workers per region. B) Using the population size and number of healthcare workers, each of the 22 regions was prioritized with regions with a large population size and high number of healthcare workers ordered first (yellow) and those with the smallest population size and number of healthcare workers ranked last (purple). The size of the point corresponds to the proportion of people over 60 years old. C) In contrast, the rankings for regions based on D) the number of confirmed SARS-CoV-2 cases (March 20, 2020 – July 30, 2021) based on the number of reported cases and confirmed COVID-19 deaths. Regions would receive doses first if they had the largest reported outbreaks (yellow) and last (purple) if they had few reported cases and deaths. The size of points indicates the number of healthcare workers per capita.

To probe this simple ranking by additionally evaluating the underlying dynamics of infection and vaccine distribution, we simulated five different vaccine distribution scenarios among the 22 regions of Madagascar assuming availability of a single batch of vaccines sufficient to vaccinate 20% of the population, and distributing doses uniformly (uniform), based on population size (pro-rata), the distribution of older individuals, defined as individuals 60+ years of age (age), reported cases (cases), and deaths (deaths). As expected based on Figure 1, there was a strong correlation between the strategies, and in all scenarios, Analamanga (the region containing the capital city) receives the largest number of vaccines (Figure 2) since it is the highest in all categories considered. We note that this initial prioritization assumes equal starting population immunity, further evaluated below.

**Figure 2:**
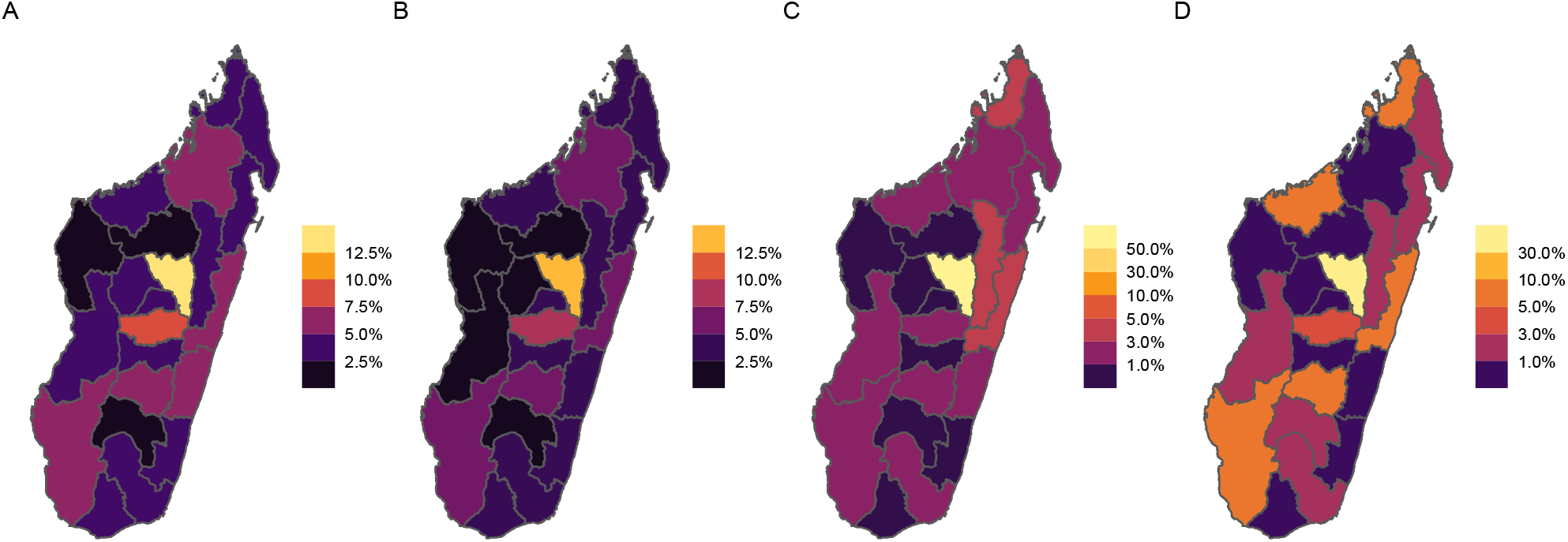
The proportion of total doses distributed by region. Assuming that the total vaccine supply is 20% of the entire population, we explored various distribution strategies. The proportion of doses per region is shown based on each prioritization scheme: (A) doses are distributed to regions based on population size (pro-rata), (B) doses are allocated based on the distribution of people aged over 60 years between the regions (age), (C) doses are distributed to regions based on the number of cases reported (cases), (D) doses are distributed to regions based on the number of deaths reported (deaths).

Overall, under the baseline scenario modeled (Figure 3A, assuming enough doses to immunize 20% of the population using a 76% efficacious vaccine with an acceptance rate of 70%, and assume that the population is fully susceptible at the start), any vaccination allocation strategy reduces the estimated number of deaths by 30-40% (Figure 3), and allocating available vaccines between regions based on the population size (pro rata) or the distribution of elderly generally outperform the other strategies (up to 10% more, Figure 3A). This is consistent across a spectrum of vaccine supply, vaccine acceptance, and speed of vaccine rollout (Figure 3B-D).The pro rata distribution of vaccines between regions or allocating doses according to the regional distribution of older people outperform distribution according to numbers of cases (or deaths) since all else equal, weighting by numbers of older individuals (which correlates with number of individuals) targets doses towards the most vulnerable [8]. As this quantity correlates with the number of Health Care Workers across regions (Figure 1B), formally modeling dose delivery does not reverse this relationship. However, if the vaccination campaign starts relatively late during the outbreak, all strategies perform equally to reduce mortality compared to a scenario without vaccination (Figure 3E) as the gains are relatively slight at this stage.

**Figure 3.**
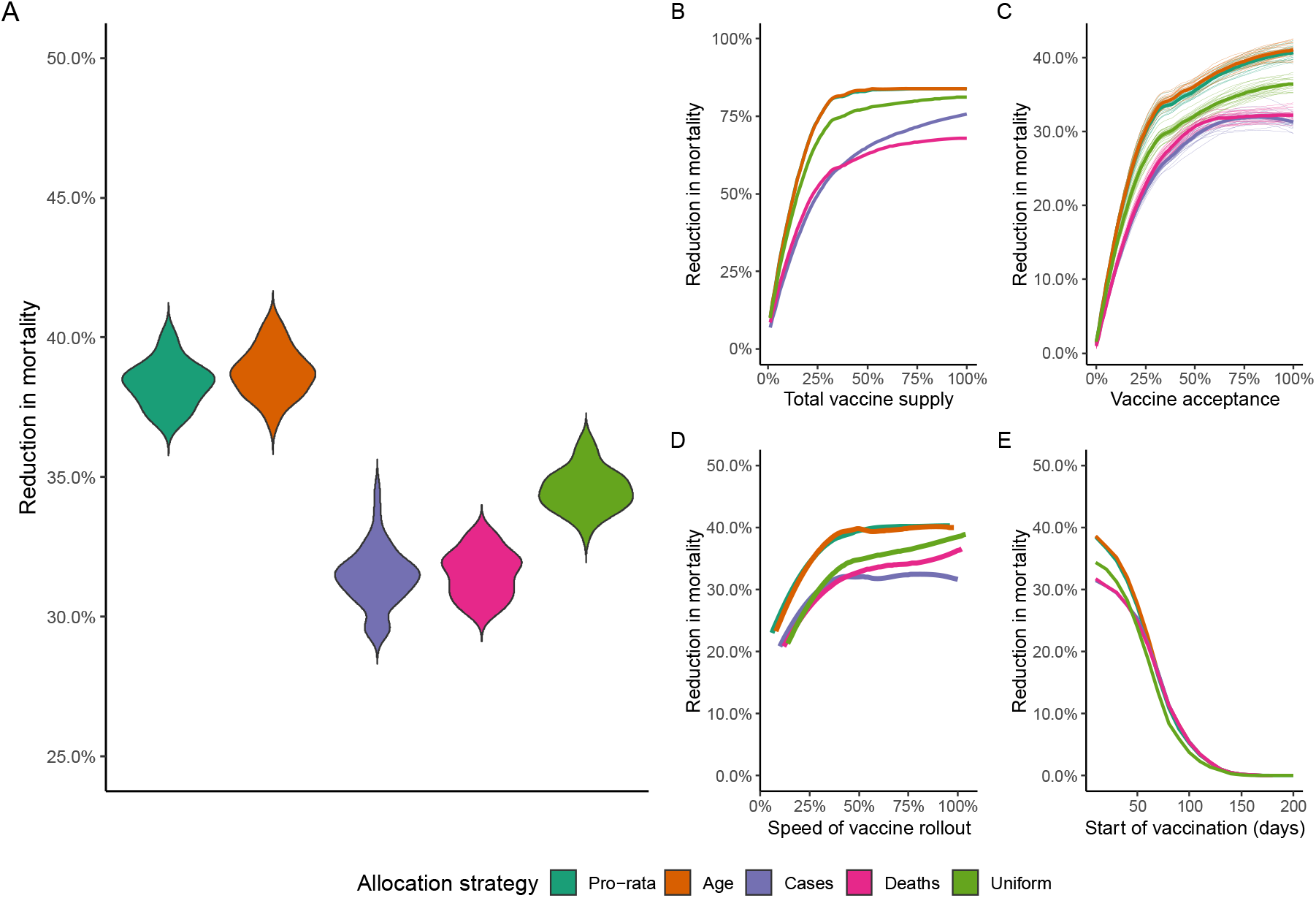
The estimated reduction in mortality for each vaccine allocation strategy. The reduction in mortality by allocation strategy for a A) stochastic simulations assuming vaccine acceptance of 70%, rollout speed where 50% of health care workers were mandated to vaccinate 20 people a day, start day of 10 days following initial seeding event, and the number of total doses equals 20% of the population; B) by varying the total vaccine supply (other assumptions assumed to be the base scenario, see Materials and Methods); C) using a range of vaccine acceptance rates; D) various roll out speeds; and E) the start date of vaccination. The median and 50 stochastic simulations are shown per sensitivity analysis

As the impact of previous waves of SARS-CoV-2 (www.covid19mg.org) on population immunity is not completely characterized [23], we evaluate two extreme scenarios: even levels of existing immunity across the country (Figure 4A, solid lines, assuming for simplicity that seropositivity and immunity are equivalent), and levels of immunity defined by reported numbers of cases (Figure 4A dashed lines). As population immunity increases (Figure 4A, x axis) the proportion of deaths averted relative to a scenario of no vaccination declines since fewer vaccines are delivered to individuals who are susceptible; however, the pro-rata and population based allocation still out-perform the other strategies. Allocation by cases and deaths performs much worse if it is assumed that immunity is distributed according to population or deaths, since doses are then targeted predominantly to the regions with lower proportions of susceptible individuals due to existing immunity (Figure 4A, pink and purple dashed lines fall fastest).

**Figure 4:**
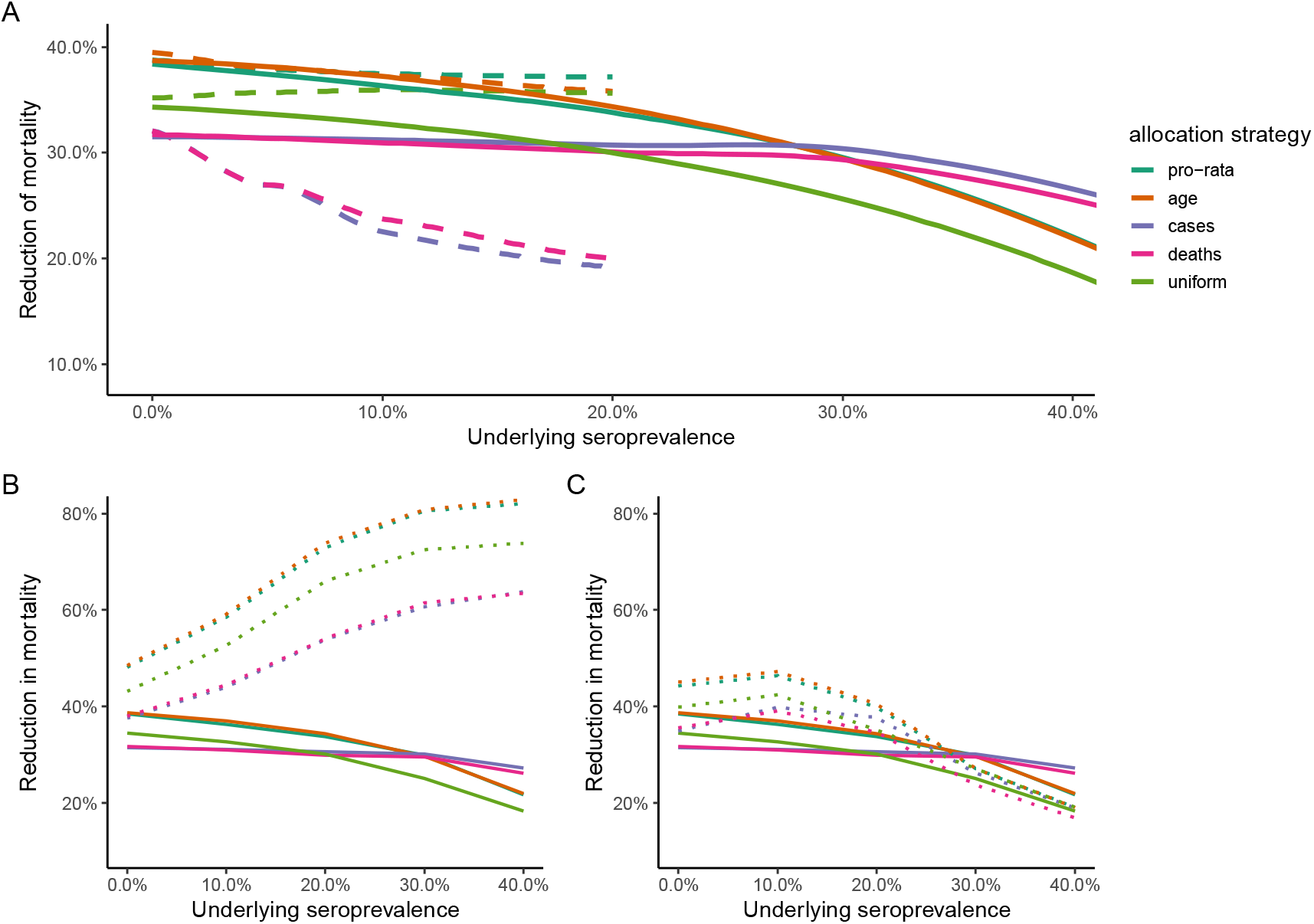
The impact of baseline population-level seropositivity on the reduction in mortality. A) The reduction in mortality by allocation strategy if population seropositivity varies between 0-40%. Two scenarios were considered: if seropositivity was distributed uniformly (solid) and by the reported number of cases (dashed lines). To avoid more than 100% seropositivity in regions with the highest number of reported cases, the case distribution maximum population-level seropositivity explored was 20% (see Supplementary Information). B) We further explored strategies where only susceptible individuals arrived at vaccination sites (dotted line) versus those with no prior information about immune status (solid line) for a range of seropositivity values (distributed uniformly). Vaccinating only susceptible individuals has the greatest reduction in mortality. C) We also investigated if testing at a vaccination site was done prior to vaccination with only seronegative individuals vaccinated (dotted line) or no prior information about immune status (solid line). These two scenarios performed similarly.

Rapid testing for sero-status (indicative of immunity) is a potential strategy to direct the deployment of doses towards those who more urgently require vaccination, as they lack prior natural immunity. We explore this by evaluating the percentage of deaths averted if only susceptible individuals are vaccinated (Figure 4B). By contrast with a scenario where vaccine doses are distributed uniformly (Figure 4B, dashed lines), when only susceptible individuals are vaccinated, the percent of deaths averted relative to the no-vaccination case increases with the proportion of the population initially immune (Figure 4B, dotted lines, gains of up to 40%), since vaccines are targeted to those who need them. However, this scenario requires all individuals to know their immune status prior to going to the health centres for vaccination (and the serological tests to be perfect), which is likely to be unrealistic. If, instead, we assume that testing occurs at health centres, thus consuming some proportion of available health care worker time, and slowing down the overall speed of vaccination (although still assuming the tests are perfect), this strategy outperforms distribution regardless of serostatus; however, initial gains in terms of deaths averted drop off as the proportion of the population immune increases (Figure 4C, dotted lines start falling at around 25% of the population immune). The gains eventually even fall below the percent of deaths averted under the uniform distribution of doses once the proportion seropositive is greater than 30%, since so many individuals will be turned away from health centres.

## Discussion

High levels of SARS-CoV-2 transmission continues to cause a global public health crisis. Mass vaccination of populations is the most effective strategy to prevent unnecessary morbidity and mortality. However, limited global vaccine supplies compel countries to prioritize among populations, and to do so in the context of an array of logistical constraints (expiration dates, healthcare worker availability, cold chains, etc.). Here, we use a stochastic age-structured model to identify dose allocation strategies that have the potential to minimize COVID-19 related deaths in Madagascar given vaccines provided by the COVAX initiative, and accounting for health care worker distribution across the country.

At a subnational scale, our results support a regional distribution strategy based on demographic parameters (population size) to allocate available doses of SARS-CoV 2 vaccines in order to achieve the highest reduction in mortality. A distribution policy based on the population size of each region is intuitively appealing as it is equitable and straightforward to implement. Indeed, pro-rata distribution of critical medicines has been used during a number of previous health crises. For example, during the 2009 H1N1 pandemic, the US Health and Human Services Pandemic Influenza Plan recommended that the different states of the USA receive pandemic vaccines in proportion to the size of its population. Additionally, researchers found that a simpler pro-rata allocation of antiviral drugs is as effective as optimal strategies targeting specific high risk groups in each region, and easier to implement [24, 25].

Given that Madagascar has now experienced multiple waves of infection (www.covid19mg.org) and that estimates of seroprevalence from blood donors in Madagascar show elevated population immunity [23], we also investigated strategies to take previous infection and thus immunity into account. Unsurprisingly, vaccinating only seronegative individuals allows for doses to be reallocated and expand protection to a larger population [8, 26, 27]. However, we note that assessing seropositivity via rapid testing at health facilities could slow the speed of vaccine delivery, which, in situations of high seropositivity, could reduce benefits in terms of mortality reductions. The benefit of additional testing to identify seronegative individuals must be weighed against the logistical challenges of testing, test accuracy [28], and ethical issues for the allocation of doses based on serostatus.

While allocating doses based on population size reduced mortality more effectively than allocation based on cases (or deaths), large heterogeneity in testing capacity between regions, and reporting issues have likely resulted in underestimates of the true burden of the pandemic, and, importantly, in a possibly spatially biased way. Although our analysis suggests that allocation based on size is robust to a number of assumptions about underlying population immunity, additional investigation, including analysing mortality records available in the capital city of Madagascar [29] and other regions, could improve estimates of transmission and identify communities where the pandemic has been particularly severe. Our analysis assumes similar starting dates for the outbreak in each of the 22 regions and neglects potential subnational heterogeneity in connectivity and contact within them [30]. The latter assumption leads to unrealistically rapid growth in case numbers within each region, such that estimates of reductions in mortality, although comparable, may be overly pessimistic. We also did not take into account time-varying estimates of transmission (Rt) resulting from the introduction of non-pharmaceutical interventions in Madagascar, given uncertainty around the magnitude of these effects emerging from data sparseness. Additional analyses that integrate data streams to better bound temporal and spatial variation in transmission could further elucidate how different allocation strategies would perform. Further, we only explored a single dose vaccination strategy, and did not evaluate the impact of various vaccines being distributed simultaneously, since only ChAdOx1-nCOV (Covishield™) is currently available in Madagascar. Finally, we assumed that seropositivity and immunity were equivalent, but decision making around the value of rapid tests for vaccine allocation will be shaped by their sensitivity and specificity [28], which may be population specific [31]) and requires careful evaluation in Madagascar

To conclude, it is clear that the speed of vaccine deployment will shape the burden of SARS-CoV-2. However, logistical limitations associated with healthcare worker numbers lead to inevitable limits associated with speed, complicating allocation across regions. Our analysis probes approaches of dose allocation across regions that most reduce mortality assuming vaccination occurs as fast as possible given these constraints, finding that allocation by population yields consistently high benefits. However, Madagascar, and many other countries worldwide fundamentally require access to more vaccine doses. Vaccine equity is the largest global issue of the present moment.

## Data Availability

The datasets generated and analysed during the current study are available in the author s github repository

https://www.github.com/fidyras/vaccination

## Declarations

### Ethics approval and consent to participate

Not applicable

### Consent for publication

Not applicable

### Availability of data and materials

The datasets generated and/or analysed during the current study are available in the author’s github repository, www.github.com/fidyras/vaccination

## Authors Contributions

FR, TR, BLR, CJEM, AW conceived and designed the paper, FR and TR wrote and performed the analyses. FR, BLR, CJEM, AW, MVE, BR, AHR, SR, and FMR wrote the manuscript.

## Acknowledgments

Authors would like to thank INSTAT for providing 2018 census data for Madagascar, the Ministry of Health for providing data on SARS-CoV2 cases and COVID-19 deaths in Madagascar. Authors would like to thank Malavika Rajeev for support in building the COVID-19 Madagascar dashboard (www.covid19mg.org).

---

**Figure S1:**
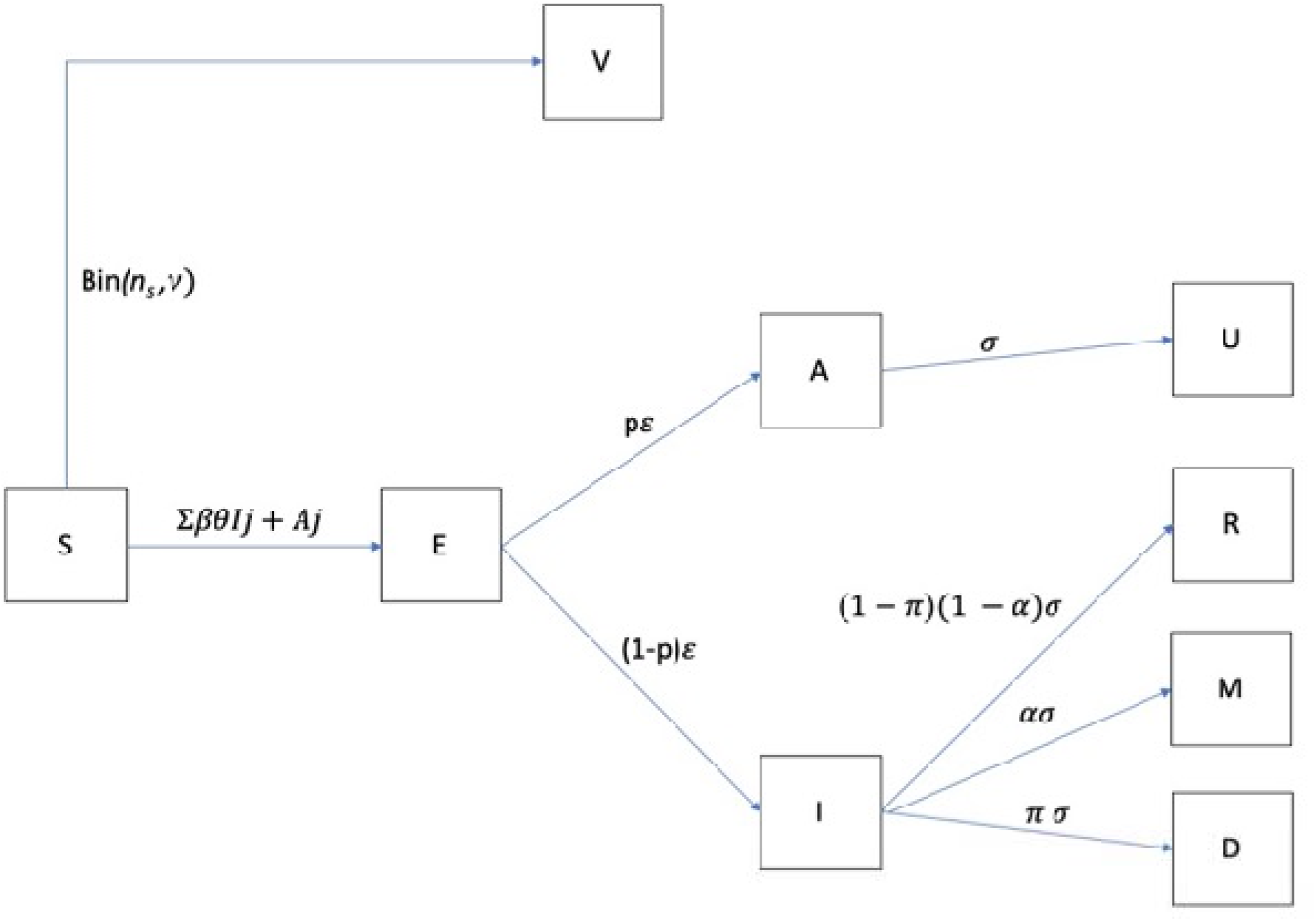
Modeling framework. The core of the model is similar to that of Roche et al. 2020. The same model has been applied to understanding the covid dynamics and the effectiveness of NPI at a national level in Madagascar (Evans et al. 2020). The stochastic simulation includes eight states: Susceptible (S), Exposed (E), Infected (symptomatic, I), Infected (Asymptomatic, A), and recovered from asymptomatic (U),recovered from symptomatic (R) infected symptomatic individuals who become severely ill (M) and infected symptomatic individuals who die (D) [17]. The population is divided into seven age classes (0-9;10-19;20-29;30-39;40-49;50-59; 60+). Susceptible individuals in an age class i (S_i_) can get the infection according to the basal transmission rate (***β***), the number of infectious (symptomatic and asymptomatic) individuals in each age class, and the contact rate among age classes. Once infected, an individual becomes infectious after an incubation period of 3 days, i.e. at a rate *ε* = 1/3 days. Infectious individuals can be symptomatic (I) with a probability of *p* (assumed to be 40%) or asymptomatic (A) otherwise. Infectious individuals (I and A) can recover at a rate *σ* = 1/5 days. Infectious symptomatic have a probability *α* to become severely ill and a probability (*π*_i_) dependent on their age class to die from the disease (D) We included vaccination into the model. The daily number of people vaccinated (*n*) depends on the number of healthcare workers and the total number of individual vaccinated is limited by the acceptance (*η*_i_). Individuals in the compartment S, E, A, U, and R can be vaccinated. However, vaccinating non-susceptible individuals would lead to vaccine wastage. Thus, we only track the number of susceptibles n_S_ that are vaccinated. Without testing, n_S_ is drawn from a binomial distribution Binomial(n, S/(S + E + A + U +R). We consider two scenarios to test for seroprevalence and thus avoid vaccine wastage (see main text). Additionally, vaccination can prioritize older individuals or distribute the doses randomly in each age class according to a multinomial distribution where the vector of probabilities are given by the frequency of the individuals in each age-class. Vaccinated individuals gain protection after a lag of 6-days according to the vaccine efficacy (*ν*) which we assumed to be at 76% to approximate the clinical vaccine efficacy against symptomatic infection seen for the ChAdOx1 nCoV-19 (AZD1222). The actual number of people gaining protection is Binomial(n_S_,*ν*).

